# The Impact for implementing Balanced Scorecard in Health Care Organizations: A Systematic Review

**DOI:** 10.1101/2021.08.05.21261666

**Authors:** Faten Amer, Sahar Hammoud, Haitham Khatatbeh, Szimonetta Lohner, Imre Boncz, Dóra Endrei

## Abstract

**Aims:** This systematic review aims to assess the impact of Balanced Scorecard (BSC) implementation at Health Care Organizations (HCOs) on Health Care Workers’ (HCWs’) satisfaction, patient satisfaction, and financial performance. Up to now, no previous systematic reviews have performed a comprehensive and rigorous methodological approach to figure out the impact of BSC implementation in HCOs.

**Methods:** This systematic review was prepared according to PRISMA guidelines. PubMed, Embase, Cochrane, and Google Scholar databases, as well as Google search engine, were inspected to find all BSC implementations at HCOs until 20 September 2020. Then the resulted articles were screened to find the implementations which measured the impact of BSC on HCWs’ satisfaction, patient satisfaction, and financial performance. Quality assessment was performed using the Standards for Reporting Implementation Studies: (StaRI) checklist.

**Results:** Out of 4031 records, 20 articles were finally included for measuring one or more of the three impact types. 17 measured the impact of BSC on patient satisfaction, 7 on HCWs’ satisfaction, and 12 on financial performance. Studies with higher quality had a higher positive impact.

**Conclusion:** This paper offers evidence to HCOs and policymakers on the benefits of implementing BSC. BSC implementations showed a positive impact on patient satisfaction and financial performance in HCOs. However, less impact was found on HCWs’ satisfaction, which should be given better consideration in future BSC implementations. High and medium-quality BSC studies were associated with higher positive impacts than low ones. BSC can be utilized as an effective tool to improve HCOs’ performance during the COVID-19 pandemic.

## 1. Introduction

In the past 3 decades, Health Care Organizations (HCOs) have been using managerial tools to enhance their performance and to achieve their targets and plans. The most known tools used to assess organizational performance are the International Organization for Standardization (ISO standards), Malcolm Baldrige National excellence model (MBNQA), European Foundation for Quality Management (EFQM) excellence model, Six Sigma, Balanced Scorecard (BSC), and other tools [1–5].

BSC differs from other managerial and performance management tools in that it can be utilized as a Performance Evaluation (PE), as well as a strategic managerial tool [6]. Many researchers suggested that the limited success of quality tools was due to the rushing into operational effectiveness with the lack of integration with the organization strategy [7–9]. The initial design of the BSC; BSC 1^st^ generation, was proposed by Norton and Kaplan in 1992. It included the evaluation of 4 perspectives: the financial, customer, internal process, and learning and growth which were steered by the organizational vision and strategy [10], see Fig (1). Strategic maps were added later in the 2nd generation of BSC to describe the cause-effect relationships between strategic objectives for each perspective [11]. In the 3^rd^ generation, destination statements, measures, and action plans were added to achieve the targets [12].

**Fig (1):**
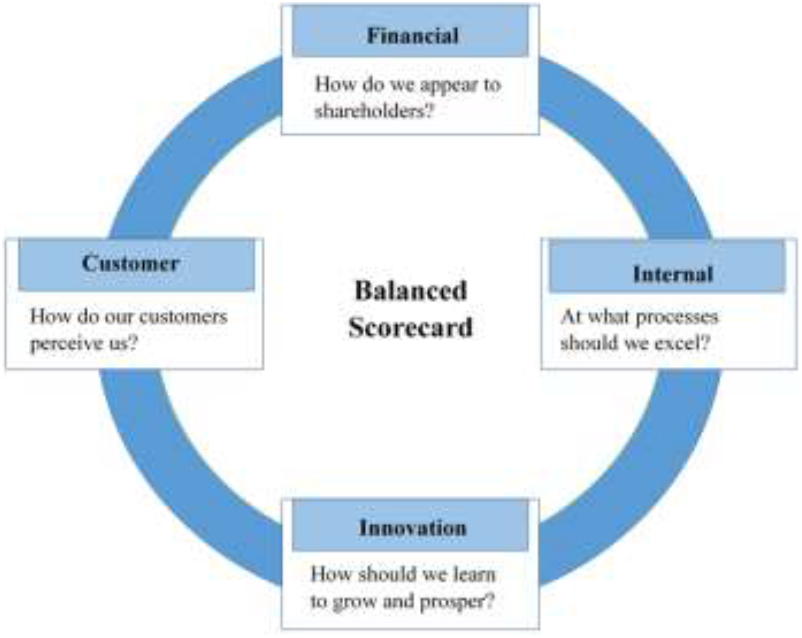
Balanced Scorecard Perspectives. [10].

Duke Children’s Hospital in the United States of America (USA) was the first health organization to implement BSC in 2000, and as a result, the hospital was able to convert 11 million dollars of losses into 4 million dollars of profits [13]. See Fig (2), which shows Duke University’s health system strategic map [14]. Since then, many hospitals around the world have implemented this tool to evaluate and develop their performance. However, the impact or the effect of BSC implementations has not yet been systematically assessed. This might be because of the varied impact types, which makes the comparison challenging [15].

**Fig (2):**
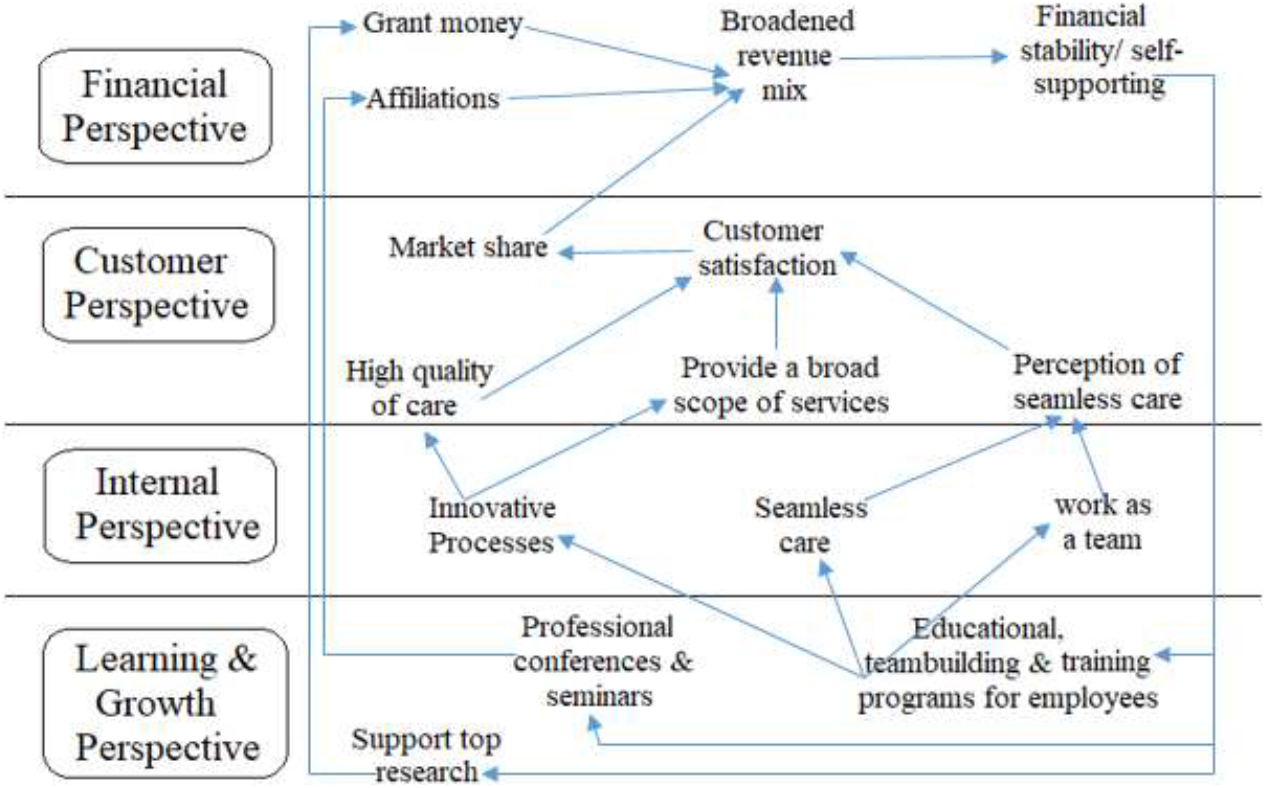
Duke University Health System Strategic Map. [14].

Coronavirus Disease 2019 (COVID-19) pandemic imposed financial burdens on countries and health care systems worldwide, as well as increased psychological stress of patients, HCWs, and the limited capacity of hospital beds during the COVID-19 pandemic [16–18]. Standard procedures and guidelines, and their delivery in full and on time were found to have an essential role in tackling COVID-19 [19]. On the other hand, the lack of standardization capability and conflicting managerial decisions were considered as dissatisfactory factors to HCWs in the pandemic [20]. BSC dimensions were suggested to have an essential role in tackling the COVID-19 pandemic by helping health care managers to mitigate its consequences on HCOs [18]. However, the effectiveness of BSC on HCOs is still not systematically evaluated. Despite the availability of systematic reviews for the impact of BSC in non-health-related fields, for example, in architecture [21] or management, marketing, and accounting fields [22], there is still limited research of BSC impact in the health care sector. Only 2 reviews discussed the impact of BSC. However, one of them did not compare the impact between the implementations or represent the impact quantitatively [15]. While the other, only mentioned few impact examples [23], indicating that none of them conducted a comprehensive or rigorous methodological approach to figure out the impact of BSC implementation in HCOs.

It is essential to evaluate the historical effectiveness of BSC implementation on HCOs’ performance before being utilized during the pandemic. Since the BSC strategic map shows that the end-results of the cause-and-effect relationships are pouring at the customer [23] or the customer and the financial perspectives [11], the objective of this systematic review was designed to assess the impact of implementing BSC at HCOs on HCWs’ satisfaction, patient satisfaction, and financial performance.

## 2. Materials and methods

This systematic review was prepared to be congruent with the 27-point checklist of the Preferred Reporting Items for Systematic Reviews and Meta-Analyses (PRISMA) checklist [24], see (S1 Appendix).

### 2.1. Eligibility criteria

The inclusion and exclusion criteria were set as shown in Table (1). Table (1) is to be placed here.

**Table (1).**
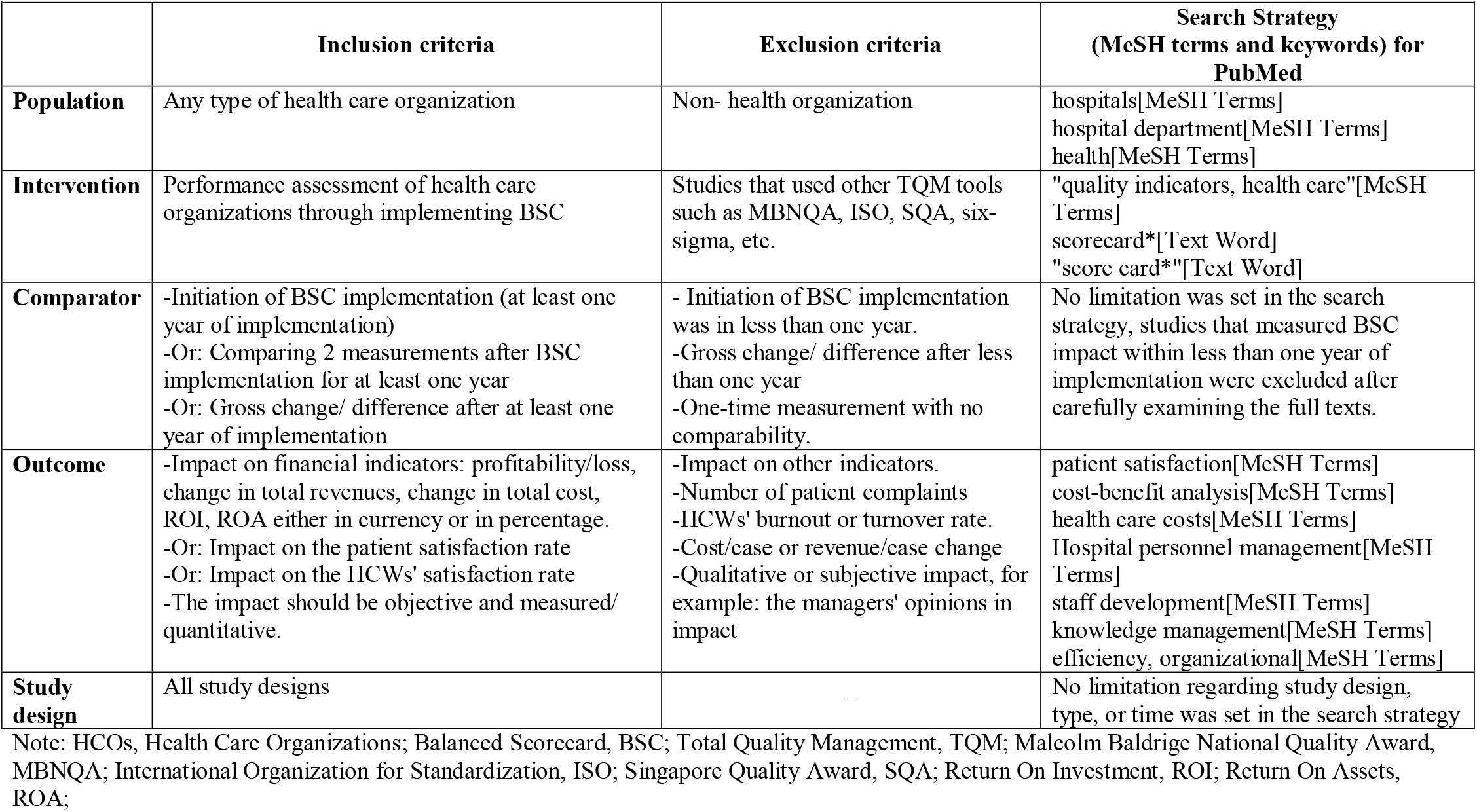
Inclusion/Exclusion Criteria and Search Strategy for PubMed.

### 2.2 Data sources, search strategy, and study selection

The search strategy was developed by the FA, SH, and SL, the first two are experts in health care management and BSC, while the third is an expert in systematic reviews and meta-analysis. The search strategy was initially developed for the PubMed database based on the PICO tool [25] which focuses on the Population, Intervention, Comparison, and Outcomes, see Table (1) using both MeSH terms and keywords. Then, the strategy was adapted for Cochrane CENTRAL, Embase, and Google Scholar databases, as per Cochrane’s recommendations [26]. To see strategies developed for these databases see (S2 Appendix). To reduce publication bias, grey literature or unpublished papers were additionally searched for in Google Scholar and the Google engine. Furthermore, we attempted to identify other potentially eligible trials or ancillary publications by searching the reference lists of included trials. The databases were searched until September 20, 2020. Afterward, FA conducted the search strategies in the electronic databases and removed the duplicates by using the EndNote X9.2 program.

The selection of eligible studies was independently performed by FA and SH. In case of disagreements, discussion after each step was made or, if necessary, SL and HK were consulted for arbitration. In the initial step of study selection, the titles and abstracts of the articles were examined to eliminate irrelevant papers. In the second step, the full texts of all potentially relevant records were carefully examined to make a final decision on in- or exclusion based on the above-mentioned criteria. Authors of studies with no available full texts or unclear impact duration were contacted to obtain further details and clarification.

### 2.3 Data collection and analysis

The following data were extracted from the final eligible studies: 1) author/s, year of publication, 2) country, 3) type of study, 4) duration of data collection 5) setting, 6) the number of health facilities, 7) the number of participants, 8) data collection tool or data sources, and 9) outcome (impact on patient satisfaction, HCWs’ satisfaction, and financial performance). Data extraction was done between January and March 2021 by FA and SH independently. The impact was either extracted directly from the studies or calculated by subtracting before and after implementation values to calculate the change; based on how each study presented its results. After that, the unification of units was performed. Next, charts plotting for each outcome were performed by FA, then reviewed by FA and SH separately. Authors were contacted if the impact measuring unit was not reported. Finally, a comparison was made to discuss the differences.

### 2.4 Quality assessment

To assess the quality of the final included papers a quality assessment was performed by FA and SH independently based on the Standards for Reporting Implementation Studies (StaRI) checklist [27]. The assessment was made for each of the included studies between March and April 2021 based on 22 items. Then the results were compared. In case of disagreement, IB and DE were consulted.

## 3. Results

### 3.1 Study Selection

The search strategy resulted in a total of 4031 records. After removing the duplicates, a total of 2985 records remained, which were screened based on their titles and abstracts. Then, irrelevant papers were excluded, so 202 papers remained. A careful examination of the resulted articles’ full texts was made, based on that 20 papers were finally included. Details of the study selection process are shown in the PRISMA flow-chart (Fig 3) and (S3 Appendix).

**Fig (3):**
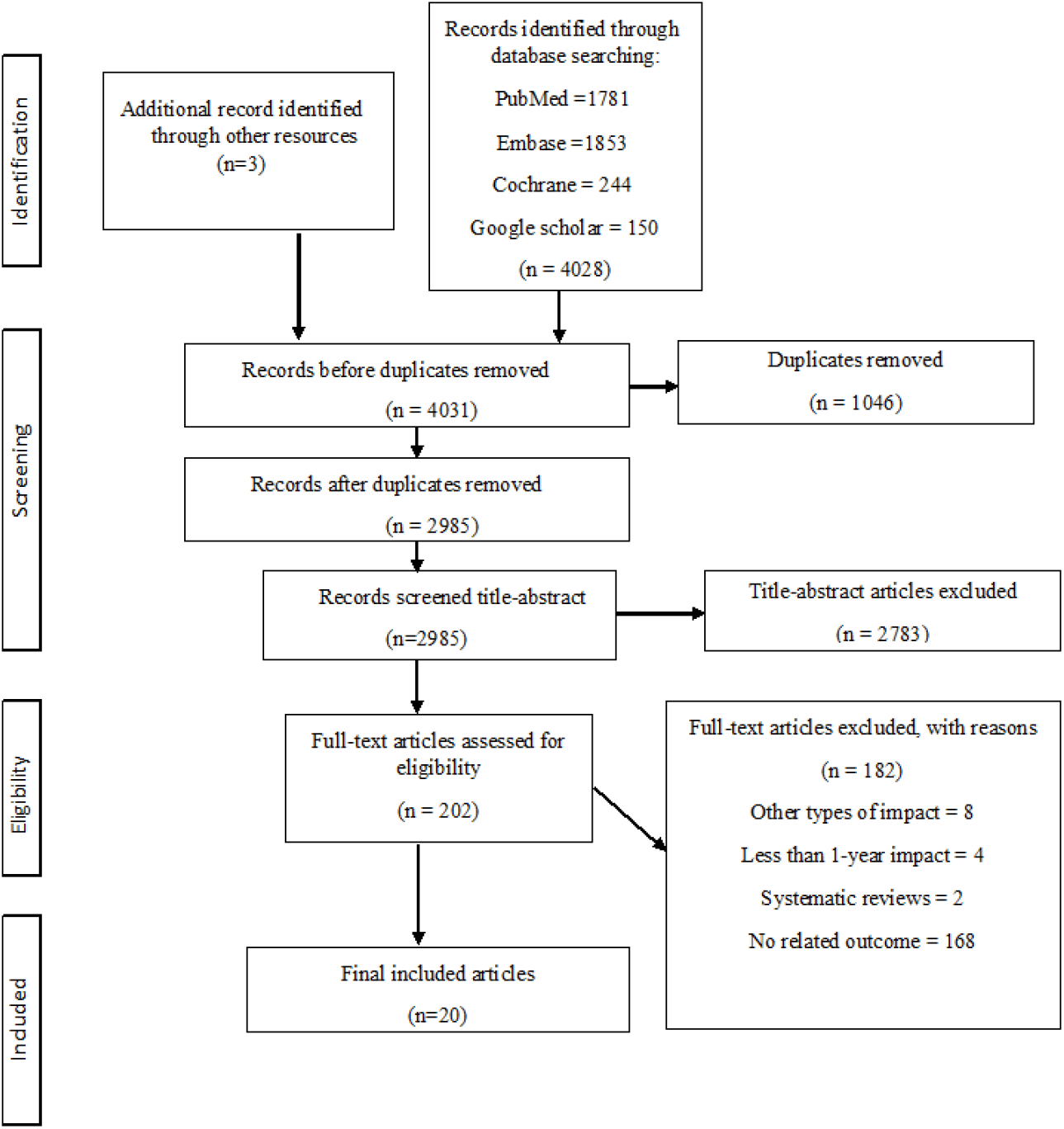
PRISMA Flow Diagram.

### 3.2 Study characteristics

The main characteristics of the included studies are shown in Table (2). The BSC implementation outcomes (impacts) are shown in (Figs 4-7) and (S4 Appendix).

**Fig (4):**
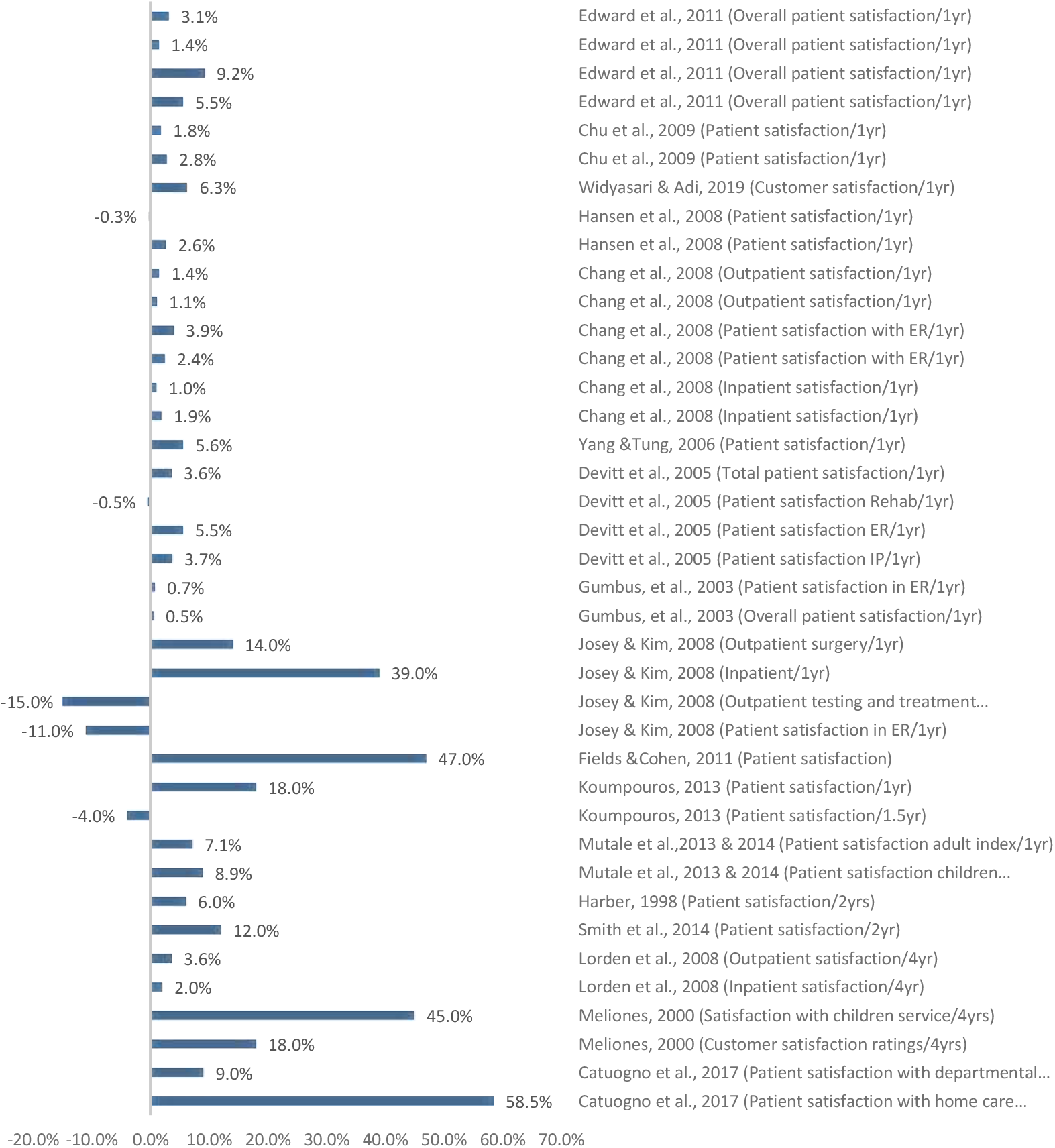
Patient Satisfaction Impact. Increase or decrease in patient satisfaction rate after BSC implementation (%)

**Fig (5):**
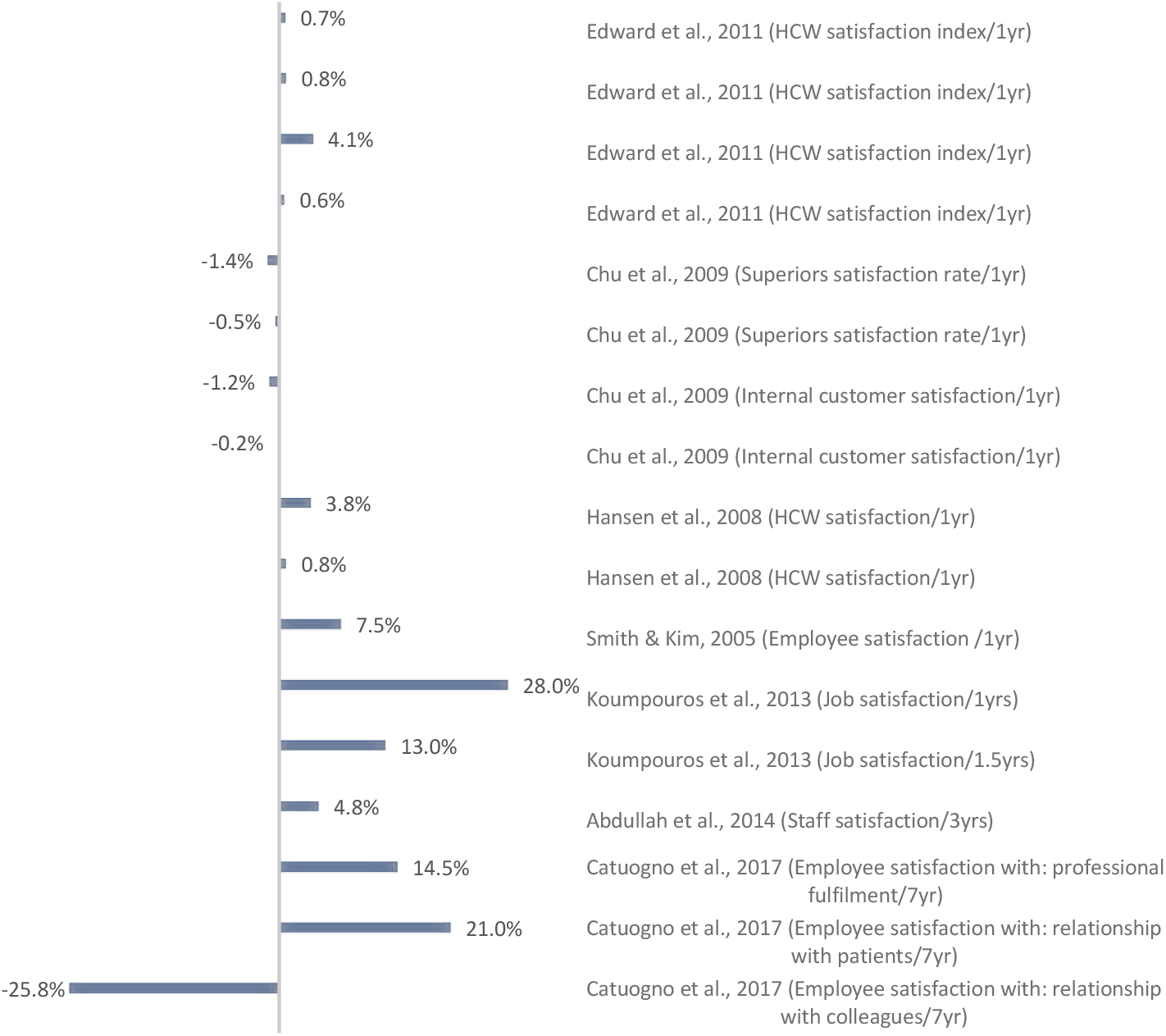
HCWs’ Satisfaction Impact. Increase or decrease in HCWs’ satisfaction rate after BSC implementation (%)

**Fig (6):**
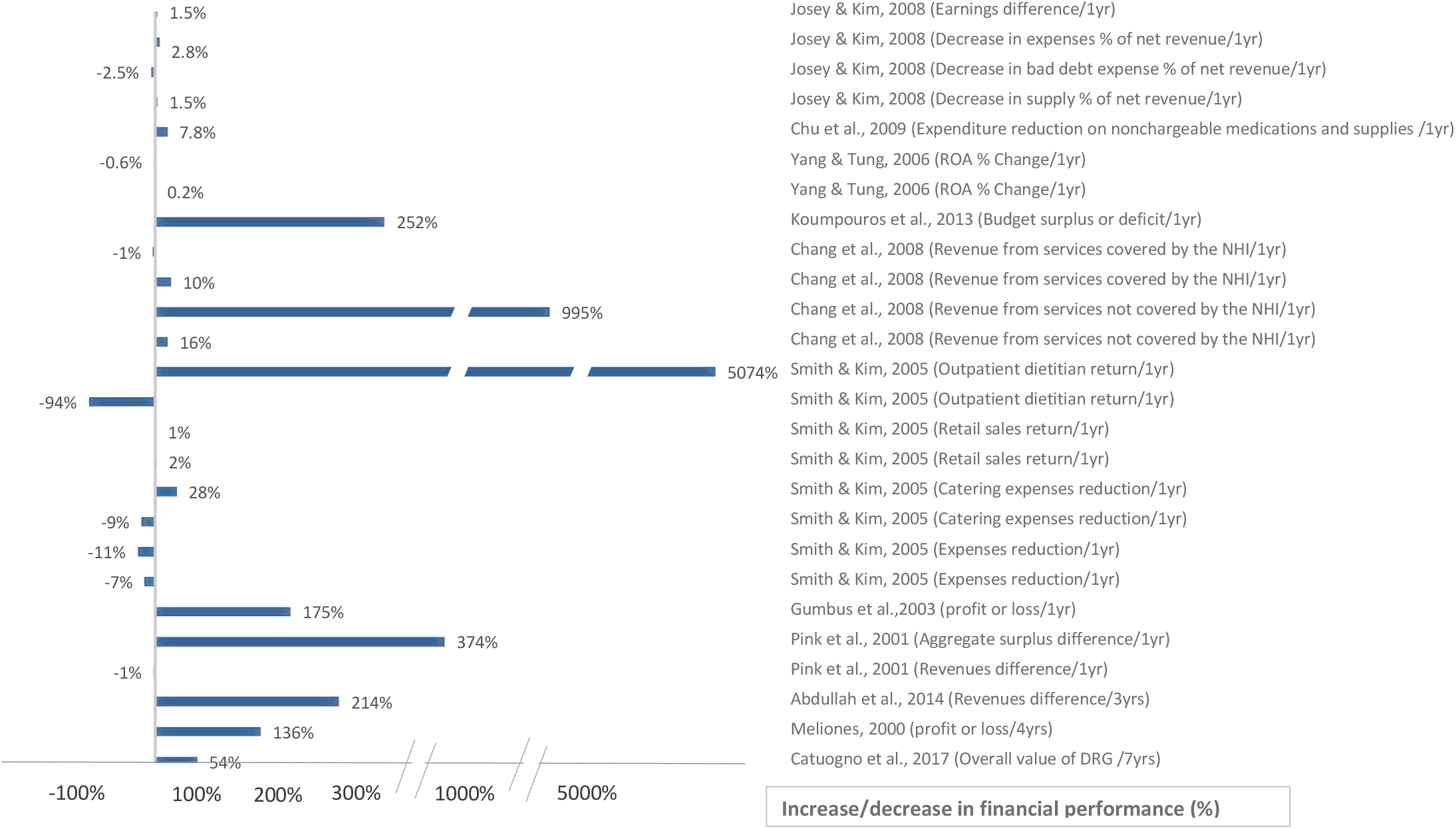
Financial Impact. Increase or decrease in financial performance after BSC implementation (%)

**Fig (7):**
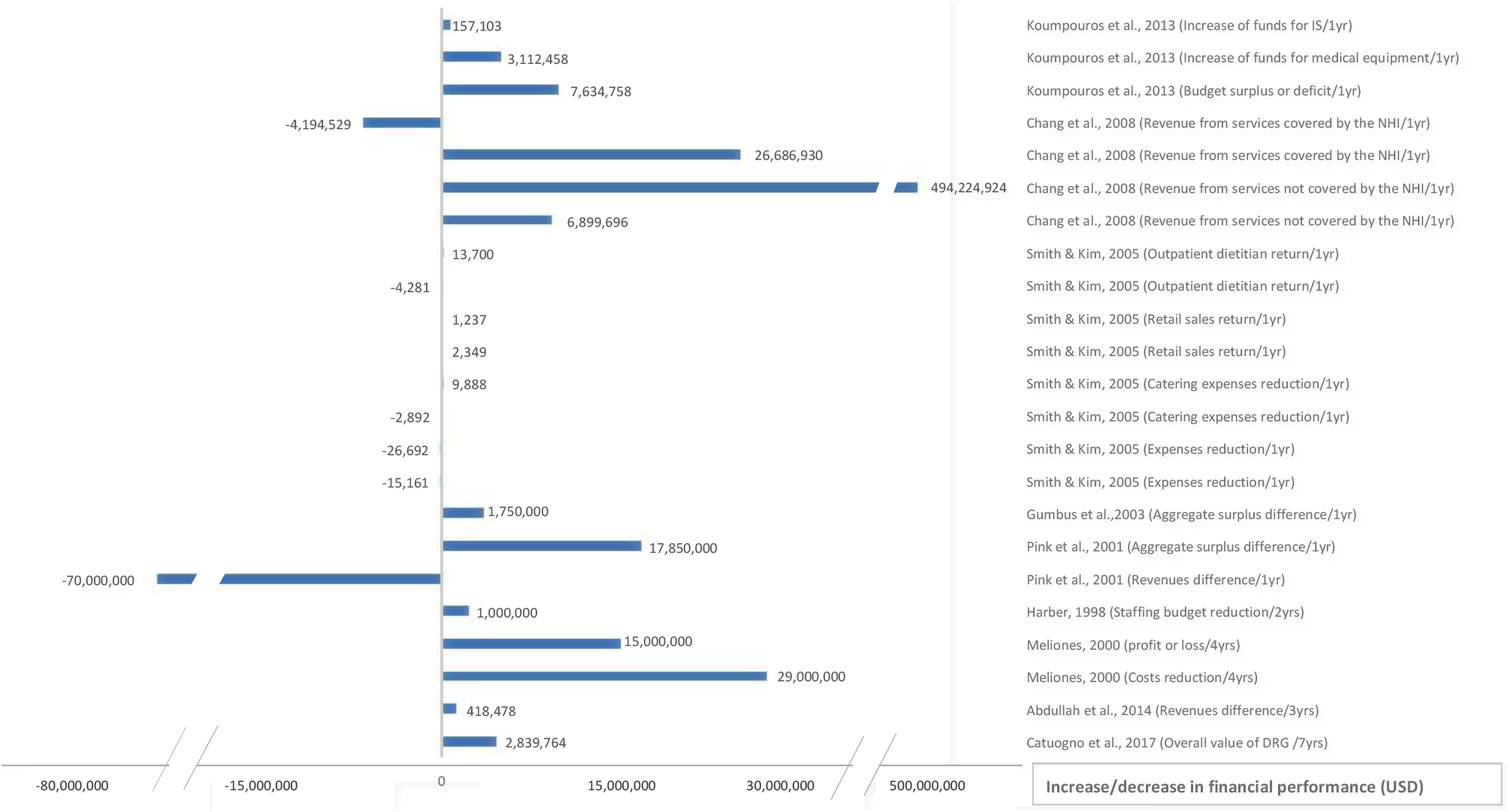
Financial Impact. Increase or decrease in financial performance after BSC implementation (USD)

#### 3.2.1 Location/ Country

Regarding the location of the implementations, 9 were implemented in North America, 2 in Europe, 1 in Africa, 7 in Asia, and 1 did not specify the location. 14 studies were performed in high-income countries, 2 in upper-middle-income countries, 1 in the lower middle, and only 2 in low-income countries.

#### 3.2.2 Setting

Out of the 20 resulted final papers, 16 were performed in hospitals or hospitals’ departments, and 4 in health facilities or clinics, see Table (2).

**Table (2).** Summary of the Final Included Studies.

| **Author/s,**<br>**year of**<br>**publication** | **Country** | **Design of**<br>**study** | **Duration of**<br>**data**<br>**collection** | **Setting** | **No. of**<br>**health**<br>**facilities** | **No. of**<br>**participants** | **Data collection tool/ data sources** |
| --- | --- | --- | --- | --- | --- | --- | --- |
| Harber,<br>1998 [46] | Canada | NR | NR | Peel Memorial<br>Hospital (Hospital in<br>general + Laboratory) | 1 | NR | NR |
| Meliones,<br>2000 [13] | The USA-North<br>Carolina | NR | 1996-2000 | Duke Children<br>Hospital | 1 | NR | NR |
| Pink et al.,<br>2001 [37] | Canada | NR | 1997-1998 | Markham Stouffville<br>hospital | 1 | NR | Surveys + hospitals reports |
| Gumbus et<br>al., 2003<br>[38] | USA | NR | 1999-2001 | Bridge port hospital | 1 | NR | The patient satisfaction measurement system |
| Smith &<br>Kim, 2005<br>[42] | USA | NR | 2001-2004 | Summa's Food &<br>Nutrition Service<br>Department at<br>Summa Health<br>System (STH & ACH<br>hospitals) | 2 | NR | Press Ganey's standard inpatient survey + audit<br>checklists |
| Devitt et al., 2005 [44] | Canada | NR | 2004-2005 | Toronto East General Hospital | 1 | NR | Data extraction from hospital records |
| Yang & Tung, 2006 [30] | Taiwan | Retrospective longitudinal | 2000-2002 | General hospitals & their supervisor agency | 21 | NR | Secondary data from the department of health + primary data structured questionnaire measuring hospitals' organizational learning and growth perspective |
| Lorden et al., 2008 [33] | NR | NR | Jan 1, 1998-June 30, 2004 (4 data collections) | Community hospital | 1 | 300 Inpatient/quarter, 700 outpatient/quarter, 227 employees (1st survey), 191 employees (2nd survey) | Employee satisfaction survey + patient satisfaction survey (via email) |
| Josey & Kim, 2008 [43] | USA-Ohio | NR | 2006 | Barberton Citizens hospital (BCH) | 1 | NR | Patient satisfaction survey |
| Chang et al., 2008 [45] | Taiwan | NR | 2001-2005 | Mackay Memorial Hospital | 1 | NR | NR |
| Hansen et al., 2008 [31] | Afghanistan | NR | July to October of (2004/2005/2006) | Health facilities | >600 | 1700 HCWs, 5800 patients-provider interaction | NR |
| Chu et al., 2009 [35] | Tawian | NR | 2004-2006 | The nursing department at a public teaching hospital in Taiwan | 1 | 13 reference nurses group | Financial data from hospital + questionnaires to executives (the weights of indicators) |
| Edward et al., 2011 [32] | Afghanistan | NR | 2004-2008 | Health facilities in Afghanistan | 700 | 1500 HCWs, 5000 patients | National health services performance Assessment + interviews with patients and HCWs |
| Fields & Cohen, 2011 [34] | USA | NR | 2009-2010 | Oregon Health and science university family medicine (Clinics) | 1 | NR | Press Ganey survey for patient satisfaction + medical records. |
| Koumpour os, 2013 [41] | Greece | NR | 18 months but not specified when exactly | General Panarcadian Hospital of Tripolis | 1 | NR | Questionnaires and interviews |
| Smith et al., 2014 [39] | Canada/ Alberta | NR | 2010-2011 (12-month trial), March 31, 2013 (results) | Hospitals in Alberta including hip and knee surgeries | 12 | NR | NR |
| Abdullah et al., 2014 [28] | Indonesia | Cross-sectional | April-December, 2013 | Cibto Mangunkusumo Hospital- Digestive endoscopy center | 1 | 76 patients | Endoscopy reports + interviews based on structured questionnaires |
| Mutale et al., 2014 [29,53] | Zambia | Cluster randomized intervention trial | 2011-2013 | Health facilities in Zambia | 12 | 96 HCWs, 429 patient interviews, 410 patient observation | A survey in facilities+ interviews with HCWs and patients + patient observation + survey with households |
| Catuogno et al., 2017 [36] | Italy | NR | 2007-2008 , & 2014-2015 | Hematology department at a Research hospital in Italy | 1 | 14 | Stakeholder satisfaction; questionnaires + care processes + hospital discharge report + charity report + research process + departmental report + economic and financial; hospital discharge database + departmental report + charity report |
| Widyasari & Adi, 2019 [40] | Indonesia | NR | During the year 2018 | Bali Mandara Hospital of Bali (Governmental hospital) | 1 | 30 | Participant observation + structured interviews + semi-structured interviews + documentation. |

Note: NR = Not Reported

#### 3.2.3 Language

Although no limitation was set based on language, all the resulted final 20 studies that measured the impact of BSC implementation were only in the English language.

#### 3.2.4 Study Designs

Only 3 studies reported their study designs clearly. 1 of them [28] referred to their design as cross-sectional studies. However, it extracted secondary data for the PE of HCOs in 3 years and compared them. While, [29] performed a cluster randomized intervention trial. It evaluated the post-intervention impact after 2 years. The 3rd study [30] reported its design as retrospective longitudinal study. The rest 17 studies did not report their design specifically. In our opinion, all the studies are longitudinal studies, which measured the PE before and after BSC implementation.

#### 3.2.5 Data collection

Variances of the used data collection tools are shown in Table (2). Only 3 studies had randomly selected HCOs, participants, or both [29,31,32]. The used indicators were clearly validated only in 6 studies [29,33–37]. Only 5 studies assessed indicators’ feasibility [34–38]. Reliability or piloting of the indicators was performed only in 3 studies [29,33,35]. Also, 5 studies only assigned weights for indicators or assessed their importance before implementation [34,35,38– 40]. Finally, only 1 study evaluated the indicators depending on more than 1 source [41], for example; patient satisfaction was evaluated from the patient’s perspective as well as the HCWs’’ perspective

#### 3.2.6 BSC generation

Resulted studies used different versions of BSC. 1st generation’s aspects; explanation and definition of perspectives and indicators, and how to measure each indicator was performed only in 7 papers [30,34,36,37,40–42], and partially in 1 paper where only customer and patient satisfaction were explained with how they were measured [43]. Also, only 5 papers specified the source for each perspective/indicator [30,35,36,41,42], while one mentioned them partially [34]. Regarding the aspects of the BSC 2nd generation, objectives for each indicator were modified by users during implementation to suit strategy, vision, mission, or goals in 5 studies [36,40,41,43,44], another 2 studies modified them partially or did not explain it sufficiently [34,38]. Strategic maps were illustrated in 6 studies [30,36,39,40,44,45], while all except 3 studies missed to display the cause-effect cascade between indicators and target/objectives [36,40,41]. Regarding the 3rd generation’s aspects, destination statements or targets within a time horizon were set in 7 studies [30,36,40–44], and only for the length of stay indicator in 1 study [13]. Strategic initiatives or action plans to achieve the targeted performance were placed in 1 study only [44].

#### 3.2.7 Impact variations

The included studies assessed different outcomes for implementing BSC. From the final eligible 20 studies, 17 measured the impact of BSC on patient satisfaction [13,29,39– 41,43,44,46,30–36,38], 7 on HCWs’ satisfaction [28,31,32,36,41,42,47], and 12 on the financial performance [13,28,47,48,30,36,38,41–43,45,46]. However, the measured variables varied among studies even for the same dependent variable, see (Figs 4-7). For example, the patient satisfaction varied from overall patient satisfaction to satisfaction of specific categories, such as adults, children, inpatients (IP), outpatients (OP), patients in the emergency room (ER), patients in rehabilitation (Rehab), or it varied according to service offered as satisfaction with home care services and with departmental services. Regarding the HCWs’ satisfaction variable, the targeted group title varied from staff, employee to HCWs, or the HCWs’ satisfaction type varied from internal customer satisfaction to job satisfaction or superiors’ satisfaction. However, the financial variable had the greatest variation; from the reduction in costs, expenditures, HCWs’ budget, expenses, catering expenses, expenses/net revenues, bad debt expenses per net revenue, supply per net revenue, to increase in: different types of revenues, return, profits, aggregate surplus, funds, the value of Drug-Related Groups (DRG), and Return On Assets (ROA).

Moreover, the unit used for financial impact assessment differed among studies. For example, all studies used currencies for assessment, where these currencies also varied between studies, except few articles which used a percentage out of 100 [30,43,47]. For the impact on patients and HCWs’ satisfaction, most articles used score assessment out of 100, except few articles which performed the assessment based on 4 or 5 points Likert scale [28,36,42]. To make the comparison of the financial impact easier, all currencies were converted to United States Dollar (USD). The authors of 1 study were contacted since they did not report the currency [36]. As a result, (Figs 6 and 7) resulted, one for the impact in currencies, and the other for the impact in percentage. While for the patients and HCWs’ satisfaction comparison, all Likert scales were converted to scores out of 100%.

Moreover, most of the studies did not specify the statistical central tendency measures used for assessments. However, a few specified using the mean to measure patient satisfaction [33] and HCWs’ satisfaction [28], while only 1 study specified using the median to measure HCWs’ satisfaction and patient satisfaction [31]. Moreover, only 2 studies mentioned testing the significance of the impact or the difference before and after the use of BSC, the first was significant in patient satisfaction [33], the second was insignificant in HCWs’ satisfaction [28]. Anyhow, all studies were included in this systematic review since significance analysis was not available in all. However, in our analysis, the magnitude of percentage change was taken into consideration.

#### 3.2.8 Time until measuring the outcome

The period between BSC implementation and measuring the impact also varied across the included studies. All articles reported results based on 1 year of implementation, except a few which reported the results of 1.5 years [41], 2 years [39,49], 3 years [28], 4 years [13,33], and 7 years [36]. For an objective comparison and to avoid bias, we reported the time between implementing BSC and assessing its impact. Due to the previously mentioned variations of measured variables, time until outcome measurement, and differences of data collection tool or data sources, see Table (2), the authors decided that conducting a meta-analysis would not lead to meaningful results, and a comparison of the impact was performed using the bar charts, see (Figs 4-7).

### 3.3 The Impact of BSC Implementation

The impacts in the 02 included studies were specified as the following:

#### 3.3.1 Impact on patient satisfaction

Using 39 measures, the impact of BSC on patient satisfaction was evaluated in 17 studies. The patient satisfaction increased after implementing BSC, since 34 measures were affected positively. Whereas, five measures were affected negatively, as the patient satisfaction unexpectedly decreased, see (Fig 4).

#### 3.3.2 Impact on HCWs’ satisfaction

The impact of BSC on HCWs’ satisfaction was evaluated in 7 studies, which contained 17 measures. 12 measures reflected a positive impact, and 5 reflected a negative impact, see (Fig 5).

#### 3.3.3 Impact on financial performance

The 12 included studies reported the financial impact either in currency or in percentage or both. These studies used 30 measures, 23 of the measures were reported in currency, and seven were in percentage. However, out of the currency measures,19 measures were presented in the papers as a monetary value before and after implementation, so the percentage change for them was possible to be calculated by us, so a total of 26 measures were plotted in percentage, see (Fig 7). But, for the remaining four currency measures presented in three studies [13,41,46], the available information was only for the final value, so it was impossible to calculate the percentage for them. As a result, 2 graphs of the financial impact were designed. (Fig 6) shows the impact in percentage, 11 studies were applicable to it, with 26 measures. While (Fig 7) shows the monetary impact in currency, where 9 studies were applicable, with 23 measures.

### 3.4 Quality assessment

Quality assessment was performed for all resulted studies using StaRI checklist. See (S5 Appendix).

## 4. Discussion

### 4.1. Discussion of the main results

This systematic review aimed at finding all the studies which measured the impact of BSC implementation on 3 variables: HCWs’ satisfaction, patient satisfaction, and the financial performance at HCOs. The impact was analyzed as the following:

#### 4.1.1. Impact on patient satisfaction

It was noticed that the studies which contained high negative impact had a low-quality assessment, except for 1 study which had a high-quality assessment and high negative impact [41], but the authors of this study explained that patient feedback revealed that patient satisfaction became lower in the 18th month due to the technical work and rearrangement of clinics noise. This explanation can be re-assured by noticing that the same impact at this study was positive (18% higher) in the first year, then it started to deteriorate in the following 6 months. However, studies that had either medium quality or high quality generally had a positive impact on patient satisfaction. Another study that had the highest quality score according to the StaRI checklist was found to have the highest positive impact on patient satisfaction [36]. This could be referred either to the lack of proper implementation of BSC including randomization, indicators’ selection, or BSC generation aspects implementation or due to the lack of sufficient reporting at this study.

#### 4.1.2 Impact on HCWs’ satisfaction

It was noticed that 4 out of the 5 negative measures of impact were mildly negative or close to zero, and were referred to in 1 study [35]. This can be referred due to the unavailability of objectives that suit the organization’s strategy, causal effects, or action plans in this study. This deficiency may have imposed a drastic effect on the final results. However, the fifth negative measure was found in a high-quality study [36]. In which, employee satisfaction with relationships with colleagues decreased by 25.7% after 7 years of BSC implementation. But, the same study found another 2 positive impacts for BSC implementation: employee satisfaction with relationship with patients, and employee satisfaction with professional fulfillment. These measures increased by 21% and 14.5%, respectively.

#### 4.1.3 Impact on financial performance

In currency, it was noticed that all high-quality studies had a positive impact for applying BSC on the financial impact in currency. Medium quality studies had a mix of positive and negative impacts. However, all negative impacts were mild; thousands of dollars compared to the millions of dollars in the positive impact. Only 1 study from the medium-quality studies had a negative impact in millions [30]; in which the revenues from the services covered by the National Health Insurance (NHI) decreased by USD 4.1 M. However, in the same study and the same year, the revenues from the services not covered by the NHI increased by USD 494 M, which was the highest positive impact for BSC in all studies included at this systematic review.

For the low-quality studies, all studies showed a positive impact in millions of dollars, except 1 study [37], in which the revenues decreased by USD 70 M. However, analyzing the decrease by percentage, shows that this decrease only represents a 1% decrease in revenues compared to the previous year. Also, another measure in the same study shows a positive impact in percentage. This will be analyzed further in the next paragraph.

In percentage, it was noticed that all high-quality studies had a positive impact in percentage for applying BSC on financial performance. Medium and low-quality studies had a mix of positive and negative impacts. However, first of all, these negative impacts were accompanied by positive financial impacts in the same study. For example, the above study shows that the revenues decreased by 1%, while aggregate surplus increased by 374% [37]. Secondly, the negative impact in percentage was very low in all studies compared to the positive impact at the same study except in 1, where the outpatient dietitian return decreased by 94% [42]. However, it increased by 5074% in the subsequent year, which represented the highest positive impact on the financial aspect in percentage.

### 4.2 Agreements and disagreements with other studies or reviews

Overall, a remarkable positive impact has been noticed for applying BSC on patient satisfaction and financial performance, this finding is compatible with another study which listed examples for applying BSC at HCOs in high-income countries [15], and a second study which listed examples for implementing BSC in general [23]. Also, the results are coherent with a systematic review that reviewed BSC’s impact in business, management, and accounting fields [22]. Moreover, in this study, a positive effect has been noticed for BSC on HCWs’ satisfaction, but with a less remarkable impact. One probable explanation for this can be referred to the lack of managerial engagement with the non-managerial HCWs upon BSC implementation, or their misunderstanding of BSC advantages, or their fear of BSC use consequences, such as to be built on for the responsibility and accountability. As a result, this may lead to cooperation resistance by the HCWs, and will also lead to lowering their satisfaction rate. However, future researchers should be considering when applying BSC on how to increase the HCWs’ satisfaction rate. One study revealed that employees did not have incentives or motives to participate in BSC since they were permanent employees, also the HCWs above 40 negatively influenced creativity and productivity upon BSC implementation [41]. This challenge was also referred to by other researchers who mentioned that in some health settings, there were major deficiencies of qualified personnel and significant issues with health care HCWs’ aging [50]. However, they have suggested that highly-ranking HCWs’ qualifications in the learning and growth perspective, will eventually generate motivation for HCWs and will resolve this issue. Other suggested ideas to solve this problem were to create an open environment for learning and growth and to encourage active communication with HCWs to ensure the successful implementation of BSC. Other research encouraged senior management commitment along with the involvement of non-managerial HCWs and clear articulation of benefits and relevancy to clinicians [51]. This is also coherent with other systematic review findings [52], which realized that health care professionals’ attitudes towards accreditation were negative and skeptical because of concerns about its impact on the quality of health care services and its cost. They suggested that health care professionals, especially physicians, have to be educated on the potential benefits of accreditation.

### 4.3 Implication during the COVID-19 pandemic

Patient satisfaction, financial performance, and HCWs’ satisfaction were severely affected during the COVID-19 pandemic [18]. Reasons for that can be referred to the psychological stress of patients and HCWs, and the limited capacity of hospital beds during the pandemic [16,17]. This paper offers evidence on the benefits of implementing BSC at HCOs on these dimensions. The implication of BSC HCOs during the pandemic may play a vital role in reducing the pandemic’s negative consequences on HCOs’ performance. This is compatible with the recommendations of other studies which emphasized the important role of the PE and managerial tools at HCOs during the pandemic [18–20].

### 4.4 Strengths and weaknesses

This paper has several strengths, according to our knowledge, this was the first paper to analyze all the studies that have measured the impact of BSC on patient satisfaction, HCWs’ satisfaction, and financial performance in HCOs. It also found a positive impact for applying BSC in HCOs, especially on patient satisfaction and financial performance. However, it suggested a need for a wider emphasis on the HCWs’ role when implementing BSC, as their satisfaction rate showed slightly positive, almost zero, or small negative scores in most studies. The 3 aspects of impact this research concentrated on, are considered the last destination for impact in the strategic maps and the causal effects at most BSC articles. Moreover, this paper shows that high-quality implementations and studies were mostly linked with higher positive results. Finally, unlike other BSC reviews [15,23] which included biobanks, pharmacies, laboratories, radiology, medical colleges in HCOs definition, this review limited the definition to the organizations which offer the 1ry, 2ry, or 3ry health care services. This strategy leads to homogeneity of the resulted studies and more valid comparisons among the results.

Nevertheless, this paper has some limitations. First, it focused on the impact of BSC at the 3 chosen indicators only. Whereas, impacts on other types of indicators were not studied. This was due to the vast variations of indicators’ types, which would have if not narrowly specified lead to analysis challenge. Second, medium and low-quality studies were included due to the lack of high-quality studies. However, this was taken into consideration when the impact was compared and analyzed. Third, no meta-analysis was applicable for this systematic review, due to the heterogeneity of studies regarding their data collection tool and the enormous variation in the types of indicators. Anyhow, the later variation was clarified in the charts, also the data collection tool was specified for each study. Moreover, only studies that measured impact after at least 1 year of implementation were included. Finally, it is important to mention that the impact comparability is likely more rational for patient satisfaction and HCWs’ satisfaction than for financial performance. Due to the feasibility to compare all studies based on a percentage out of 100 score for the first two variables. While the change of financial performance based on currency is harder to compare among studies due to its dependence on other factors such as the HCOs’ size or the number of health facilities included in the study, etc. These factors may have played as confounding factors on the amount of financial impact. Also, comparing financial performance based on the percentage change does not imply the real performance change, because the initial magnitude before BSC implementation could be so small, which would have shown a vast increase in percentage even if a small increase in the amount occurred. So, the financial performance comparison among articles gave only a general idea regarding the impact differences, while considering other factors in the analysis was important too, also using both percentage and currency in the analysis would give a better holistic idea. Finally, due to the difference of currency used among studies, the currency was converted to USD. However, for studies that did not specify the rate at the time of the study, the current rate was used in calculations.

## 5. Conclusions

In conclusion, the implementation of BSC positively influences patient satisfaction and financial performance in HCOs. However, future researchers should have more attention to HCWs’ satisfaction and engagement at BSC implementations. This paper offers evidence to HCOs and policymakers on the benefits of implementing BSC in health care. Moreover, BSC may play an effective role in improving HCOs’ performance during the COVID-19 pandemic.

## Data Availability

The data underlying this article are available in the article and in its online supplementary material.

## 4 Acknowledgements

We would like to thank Dr. Vahideh Moghaddam, Diêgo Andrade, and Wang Zhe for their help in translating and data extraction of Persian, Spanish, and Chinese articles respectively. Also, we would like to thank Dr. Duha Shellah for contributing to revising the manuscript.

## 7. Appendices

**S1 Appendix. PRISMA checklist**. 27-point checklist of the Preferred Reporting Items for Systematic Reviews and Meta-Analyses (PRISMA) checklist (DOCX).

**S2 Appendix**. Search strategies in PubMed, Embase, Cochrane, and Google scholar (XLSX).

**S3 Appendix**. Title-abstract screening, and full text screening (XLSX).

**S4 Appendix**. Data extraction of impact and calculations (XLSX).

**S5 Appendix**. RoB and quality assessment (XLSX).

